# “I prefer a doctor on camera than none at all”: Participants’ Perspectives on the Implementation of Integrated Telehealth and Community Health Workers Services to Improve Access to Healthcare in Rural Communities

**DOI:** 10.1101/2024.11.13.24317260

**Authors:** Jose G. Perez-Ramos, Scott McIntosh, Joselyn Wei-Chen, Jessica Alicea-Vellon, Carlos Rodriguez-Diaz

## Abstract

**Background:** Rural and isolated communities, such as Culebra, Puerto Rico, face significant healthcare challenges due to geographic isolation, limited medical resources, and socioeconomic disadvantages. Chronic diseases, particularly diabetes and hypertension, are prevalent and contribute to poor health outcomes. Telehealth services (THS) and community health workers (CHWs) have been identified as effective interventions for improving healthcare access in underserved areas. This study aimed to explore the community perceptions and attitudes towards integrating THS with the support of CHWs to enhance chronic disease management in Culebra.

**Methods:** A Qualitative Research approach involved 20 patients from Culebra’s Federally Qualified Health Center (FQHC). Semi-structured interviews were conducted, guided by the Socio-Ecological Model (SEM), to assess community perspectives toward THS and CHWs at individual, interpersonal, community, and societal levels. Thematic analysis revealed three primary themes: social determinants of health, the role of CHWs, and the benefits and challenges of THS.

**Results:** Findings indicate that CHWs play a critical role in improving healthcare access by assisting with medical appointments, medication management, and providing emotional support. THS was recognized for its potential to reduce transportation barriers and improve continuity of care, though concerns regarding convenience, technology access, and data privacy persist. The integration of CHWs and THS was viewed positively as a solution to healthcare access challenges, with participants expressing trust in this model for managing chronic diseases.

**Discussion:** This study highlights the need for further development of THS infrastructure and CHW support to ensure equitable healthcare access in rural areas like Culebra. The combination of THS and CHWs shows promise for reducing healthcare disparities and improving outcomes for isolated populations. Future efforts should focus on systemic changes to address broader social determinants of health, fostering sustainable and effective healthcare delivery in underserved communities.

## BACKGROUND

In the United States (US), one in five communities represents a rural or isolated area.(1) Rural and isolated communities face numerous healthcare challenges, typically as consequences of having limited access to healthcare due to their lower socioeconomic status and geographic location relative the services.(2, 3) The health outcomes of people living in rural communities, such as survival rate, complication frequency, and disease management, are often worse compared to people living in urban areas, resulting in a significant public health crisis.(4) The same factors impact the health and healthcare of persons in Latinx communities, including a higher prevalence of chronic conditions such as diabetes (10.3% in Hispanic communities compared to 8.5% in non-Hispanic white communities) among adults aged 18 years or older.(5) These disparities are even more pronounced for people living in Puerto Rico, who experience the worst health outcomes compared to the general US population, having an estimated diabetes prevalence of 20.1% among adults.(6) Puerto Rico (PR) is a territory of the US that includes many rural communities that experience further isolation by an amalgam of factors. Chronic diseases, such as diabetes and hypertension, represent pressing public health problems in many rural and isolated communities in PR. According to The Centers for Disease Control and Prevention (CDC), chronic diseases are the leading cause of death among adults in the US and PR.(7) Compared to other jurisdictions in the US, previous studies had suggested a higher prevalence of chronic diseases among people in PR; hypertension (39% vs 36%) and high cholesterol (16% vs 10%).(8)

Most people in PR (98.9%) self-identify as Latinx/Hispanic in official federal documents. PR is an archipelago with three inhabited islands; most people live in the “big island” of Puerto Rico, and the two populated island municipalities are Culebra and Vieques. Culebra is the smallest of the inhabited islands, located 17 miles east of the island of PR with an extension of 11.62 mi^2^. The island of Culebra is the least populous municipality of PR, with a population of 1,792. Among its residents, the median age is 44.6 years, 23.5% live under the poverty level, 56.7% are unemployed, and 13.5% don’t have healthcare coverage.(9) The estimated prevalence of diabetes and hypertension were 15.1% and 39.2%, respectively, in Culebra (2019).(10) The people of Culebra have historically faced numerous challenges accessing healthcare services.(11) Geographic isolation, scarce medical resources, and a shortage of healthcare providers all affect the consistent medical care, particularly for those with chronic diseases. They rely on the health services on the main island, already strained by the lack of enough providers due to migration, causing long waiting times for service.(12) The lack of reliable transportation to the main island exacerbates these barriers to access to care. (13–15) These challenges are social determinants of health (SDoH) that result in significant health inequities affecting the lives of island residents.

Healthcare responses to the COVID-19 pandemic have demonstrated that Telehealth services (THS) and the deployment of community health workers (CHWs) increase access to care and improve health outcomes, particularly in rural areas that were previously underserved.(16) However, THS has not been equitably available, and rural communities such as those in PR have yet to benefit from this opportunity for increased access.(17) The healthcare disparities and challenges associated with chronic disease management faced by Culebra or similar communities have not been fully addressed, particularly with the integration of CHWs and THS in improving the health outcomes of rural and isolated communities.

Considering the opportunity to provide evidence-based services to improve access to care services for Latinx populations in isolated communities, this study aimed to explore the community perceptions and attitudes toward integrating THS with the support of CHWs to improve healthcare access for people with chronic diseases in isolated areas such as the island of Culebra, PR.

## METHODS

### Research Team

This research was implemented by an established Community-Academic Partnership (CAP) between a research academic institution and HealthproMed Inc., the only Federally Qualified Health Center (FQHC) in Culebra. The CAP identified the need to understand how the community perceived the potential implementation of two evidence-based interventions (EBI): 1) CHWs assisted 2) THS for patients with a specialist referral to manage their chronic disease (e.g., diabetes and hypertension). The research team integrated the Socio-Ecological Model (SEM) as a theoretical framework to explore how the community perspectives were related within and across different and multiple levels. With this approach, findings would support the comprehensive implementation of these integrated interventions.(18)

The research team comprised a multidisciplinary group of professionals, including a trained and certified community health worker in Culebra, FQHC personnel, including a nurse and health specialist provider, and the study team located in Rochester, NY. The team’s collective efforts aim to create a culturally congruent, ethically sound research environment that contributes to the academic field and brings tangible improvements to healthcare access and better management of individuals in isolated island communities like Culebra. This study was approved by the URMC Institutional Review Board #STUDY00007257 and followed COREQ guidelines to report qualitative research.(19)

### Study Design

This study focused on ascertaining community perceptions toward THS infrastructures and the role of CHWs in addressing health access for people with chronic diseases. For this qualitative study, we recruited 20 community patients from the local FQHC in Culebra, using a purposive sampling methodology and direct recruitment efforts from CHWs. A total of 20 participants were recruited, which was considered sufficient to provide adequate information power. Information Power approach facilitates the justification for this smaller sample size by having the study with a specific aim, a focused sample, and strong interactions between participants and researchers.(20) Participants were required to be older, at least 18 years old, live in Culebra, have a chronic disease diagnosis, and be able to read the information provided in Spanish for informed consent. The exclusion criteria included non-residents, non-patients of the local FQHC, and unable to communicate in Spanish. The study was conducted in Spanish because it is the official language in PR and the primary language of most of Culebra’s residents.

An interview guide for semi-structured qualitative interviews was developed and conducted by a team member proficient in qualitative research methodologies and fluent in Spanish.(21) The interview guide was developed using the SEM framework to capture in-depth perceptions of THS and the role of CHWs in the community as an intervention to increase access to healthcare. Our research team member experienced in qualitative research methodology conducted all interviews in Spanish. The Rapid Qualitative Inquiry (RQI) method allowed for a dynamic and iterative data collection and analysis process, which supported the identification of salient themes and patterns.(22) Participants were recruited from the clinic’s patient population and were informed of their rights to discontinue participation or skip questions at any point. The interviews lasted up to an hour and were conducted in familiar community settings, such as the facilities of the FQHC and community centers, ensuring a comfortable and private environment for participants to share their experiences. Thematic analysis was conducted and supported the identification of salient topics shared by participants.(23) Themes and supportive quotes were translated and back-translated from Spanish to English by two Spanish-speaking speakers members of the research team following the Brislin Back-Translation Method to address interval validity.(24)

Participants were compensated with $10.00 cash at the end of the interviews. Additionally, they were offered refreshments in a culturally congruent manner to express gratitude for their participation. The study design prioritized participant privacy and data protection. Data were collected anonymously, and participants were encouraged not to disclose identifiable information during the interviews. However, any identifiable information to participants was removed from transcripts and data analysis. Risks were minimized by de-identifying all data in the final analysis, thus preserving participant confidentiality.

Interview audio files were initially stored on encrypted phones, transferred to encrypted computers, and uploaded into a cloud-based HIPAA-compliant software with 256-bit encryption and SSL-EV to ensure the highest level of data protection.

Table 1 presents the demographic profile of the 20 participants, who predominantly self-identified as female (n=12; 60%) and with a diverse age range (20-70 years old). Most participants shared their age, including from their 20s to their 70s. However, (n=8; 40%) of the participants declined to disclose their age. The average interview duration was 55 minutes and 42 seconds. Participants’ educational levels varied, with a slight majority having completed or exceeded high school (n=11; 55%) and a quarter having less than a high school education. Interviews were conducted between October 2022 and March 2023.

**Table 1.** Qualitative Research Logistic and Demographic Information.

| Criteria | Description |
| --- | --- |
| Gender | Male = 8 (40%)<br>Women = 12 (60%) |
| Interviews Length | 55 min 42 sec (AVERAGE) |
| Age | 20s: N=2 (10%)<br>30s: N=1 (5%)<br>40s: N=2 (10%)<br>50s: N=3 (15%)<br>60s: N=3 (15%)<br>70s: N=1 (5%)<br>Undisclosed: N=8 (40%) |
| Educational Levels | <HS: N=5 (25%)<br>=HS: N=9 (45%)<br>>HS: N=2 (10%)<br>Undisclosed: N=4 (20%) |

### Data Preparation and Analysis

Qualitative data analysis was conducted using Dedoose^®^, a qualitative analysis software that allows for direct coding.(25) Interview data were examined iteratively using thematic analysis for patterns and themes, investigating community attitudes and perceptions toward THS systems and the community health worker’s model.(23) The analysis was guided by the SEM,(18) which provided a comprehensive understanding of participants’ attitudes and perceptions toward our integrated model. The team regularly (≥5 times per week) debriefed about findings during fieldwork with partners and community members.

## RESULTS

Identified themes and subthemes were categorized across SEM domains to provide a comprehensive understanding of the factors influencing participants’ perceptions of implementing integrated, evidence- based interventions. As shown in Figure 1, the SEM domains include Individual, Interpersonal, Community, and Societal levels, each addressing three main themes with subthemes: social determinants of health, the role of CHWs, and the impact of THS services.(18) This categorization helped illustrate how these factors interact and contribute to the understanding of the community’s perspectives and attitudes toward integrating THS with the support of CHWs to improve healthcare access for people with chronic diseases on the island of Culebra. The most relevant subthemes of each theme will be discussed in the following sessions.

**Figure 1.**
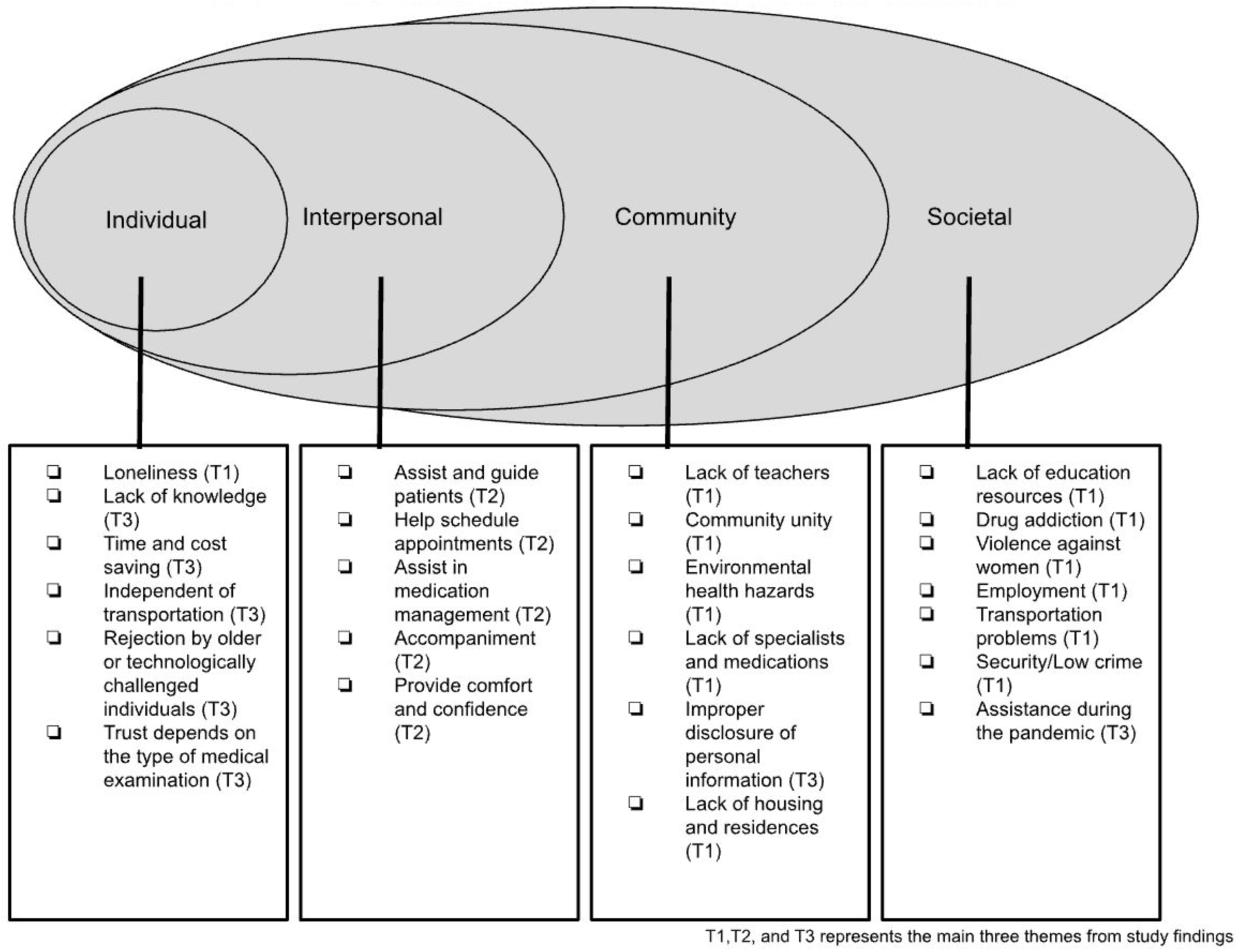
Applied Social Ecological Framework to the Thematic Analysis

### Theme 1 (T1): Social Determinants of Health

Participants highlighted several key issues affecting their community, including limited educational resources, economic instability, violence, and inadequate housing and transportation (Table 2). These factors create significant barriers to health and well-being, emphasizing the need for community-based interventions and policies to address these underlying causes of health disparities in rural and isolated areas.

**Table 2:** Theme 1 - Social Determinants of Health.

| Domains | Individual | Interpersonal | Community | Societal |
| --- | --- | --- | --- | --- |
| **Subthemes** | • Lack of education resources<br>• Loneliness<br>• Employment<br>• Lack of housing and residences<br>• Lack of medications | Employment | • Lack of education resources<br>• Employment<br>• Community unity<br>• Environmental health hazards<br>• Lack of medications | • Lack of education resources<br>• Employment<br>• Lack of housing and residences<br>• Transportation problems<br>• Safety/Low crime<br>• Community unity<br>• Environmental health hazards<br>• Lack of medications |

### Subtheme: Transportation Problems

Transportation was identified as a significant issue, with the community’s geographical isolation contributing to difficulties in accessing other areas. This affects daily life and service provision. A participant explained, “Well, transportation. Like Vieques and Culebra, we are, as you say, far from the big island, so sometimes we have a lot of problems with transportation, that’s more than most of what it affects” (IND49). This reflects societal-level challenges.

### Subtheme: Environmental Health Hazards

The improper management of sewage and wastewater was identified as a severe environmental and health concern by the Culebra community. Overflows cause a threat to the cleanliness and hygiene of the community, as expressed by one participant: “Sewage is overflowing everywhere. It is really a very strong concern.” On the other hand, participants also discussed the negative impact of exploitation- induced destruction, particularly from investors and unsustainable practices. This leads to the destruction of natural resources and cultural heritage and threatens the community’s long-term well-being. A participant stated, “Our natural resources are being destroyed. The new generations are the ones who are going to have to deal with the exploitation” (IND2). Both issues affect the community and societal levels.

### Subtheme: Lack of Medications

A consistent medication shortage was identified as a pressing health issue, particularly affecting emergency services and controlled medications. This hinders the effective management of health conditions. One participant highlighted, “There is a lot of lack of medication. In the emergency area, there is a lot of medication shortage” (IND2). This issue impacts individual, community, and societal levels.

### Subtheme: Lack of Education Resources

Participants identified a shortage in the availability and quality of educational resources, exemplified by a lack of schools and the underutilization of existing educational spaces. This lack of resources was seen as a barrier to providing adequate education within the community. For instance, one participant stated, “Everything that has to do with education, that is what we are looking for” (FG1). These insights reflect significant concerns at the individual, community, and societal levels.

### Subtheme: Loneliness in Some Individuals

Feelings of isolation and loneliness were identified as significant concerns for specific community members, particularly during times of personal crisis. Loneliness affects mental and emotional well-being. For example, one participant noted that some patients like to come to the clinic to have company “because many times they are alone, and what they want is to talk” (IND2). This reflects the individual-level impact of social isolation.

### Subtheme: Employment

A lack of diverse employment opportunities was highlighted as a significant issue, especially for the youth. The local economy offers limited career paths, forcing individuals to migrate or accept employment outside their desired professions. A participant explained, “For young people, there are not many [options out] there; what we have are the same [jobs] in a restaurant, but there is no way that if someone wants to be a lawyer, there is no way. There are not many alternatives for work” (FG1). The decline in the available labor force, mainly due to an aging population, was seen as a challenge in maintaining essential services. This sense of vulnerability affects those who are aging or ill. This issue impacts the individual, community and societal levels.

### Theme 2 (T2): The Role of Community Health Workers

Community Health Workers are vital in enhancing healthcare access and quality for individuals in underserved communities (Table 3). They provide essential support by assisting with patient education, scheduling medical appointments, managing medications, and offering companionship during medical visits. Their involvement improves health outcomes and fosters a sense of trust and comfort among patients, significantly impacting their overall healthcare experience. However, most people do not fully understand CHWs’ role, even though some might already have been in contact with CHWs before.

**Table 3:** Theme 2 – The role of Community Health Workers.

| Domains | Individual | Interpersonal | Community | Societal |
| --- | --- | --- | --- | --- |
| Subthemes | • Helps schedule appointments with specialists<br>• Assists in medication management<br>• Provides comfort and confidence | • Assist and guides the patient<br>• Helps schedule appointments with specialists<br>• Assists in medication management<br>• Trust and social/health support<br>• Provides comfort and confidence | • Provides comfort and confidence | • Provides comfort and confidence |

Therefore, it is essential for people to know what CHWs can do for them to improve their healthcare access.

### Subtheme: Assists and Guides the Patient

Participants highlighted the vital role of CHWs in providing valuable assistance and guidance. CHWs help patients understand their health conditions and learn how to manage their treatments, improving their autonomy and care. One participant noted, “They guide you, give you information so you know what these symptoms are based on, and how to take care of yourself” (FG103). Another shared, “They have taught me how to inject insulin. I have done it, but I have had several doubts. And they clear up my doubts” (IND39). These interactions enhance the interpersonal relationships between CHWs and patients, fostering trust and confidence.

### Subtheme: Helps Schedule Appointments with Specialists

CHWs play a crucial role in helping patients schedule appointments with specialists, ensuring continuous and specialized care, especially for chronic conditions. A participant explained, “If I needed such an appointment with the specialist, find me one. When I need a referral, what I do is that I go with the community health worker, we interconnect, and things seem to move a little faster; that’s why it’s important” (IND2). This assistance is essential in maintaining the continuity of care and addressing the specific health needs of patients.

### Subtheme: Assists in Medication Management

The support provided by CHWs in medication management is seen as crucial, particularly for those who struggle with maintaining their medication regimens. One participant stated simply, “They get the medicines for me” (IND59). This service is essential for ensuring that patients adhere to their prescribed treatments and manage their health conditions effectively.

### Subtheme: Trust and Social/Health Support

Participants valued the accompaniment provided by CHWs when accessing medical facilities. This support offers comfort and confidence, particularly when patients need to access laboratory services or return home. One participant shared, “And I have a lot of confidence in her when I have to go to the laboratory. She waits for me patiently. She takes me home” (IND59). This accompaniment helps reduce the stress and anxiety associated with medical appointments, contributing to better health outcomes.

### Subtheme: Provides Comfort and Confidence

CHWs are recognized for their attentive and considerate care, which instills comfort and confidence in patients. As one participant expressed, “They gave me comfort” (IND49). This emotional support is essential to the healthcare experience, helping patients feel more secure and supported in managing their health.

By offering these services, CHWs play an integral role in improving healthcare delivery and outcomes, particularly in underserved communities. Their work addresses both the practical and emotional needs of patients, fostering a more supportive and effective healthcare environment.

### Theme 3 (T3): Telehealth Services

Participants expressed different perspectives towards the THS Services in providing healthcare access in rural and underserved communities for chronic disease management (Table 4). Some participants believed that THS could offer numerous benefits, including reducing the need for transportation, saving time and costs, and ensuring continuity of care when in-person medical visit options aren’t available or accessible. However, THS also faces challenges such as perceived inconvenience compared to in-person visits, lack of knowledge among users, and concerns about data privacy. Despite these challenges, THS was seen as a beneficial alternative when traditional healthcare options are inaccessible, enhancing overall healthcare experiences by offering peace of mind and improved communication between patients and providers.

**Table 4:** Theme 3 – Telehealth Services.

| Domains | Individual | Interpersonal | Community | Societal |
| --- | --- | --- | --- | --- |
| Subthemes | • A good alternative when no other option is available<br>• Lack of knowledge<br>• Time and cost saving<br>• Possible improper disclosure of personal information<br>• Rejection by older or technologically challenged individuals<br>• Trust depends on the type of medical examination<br>• Peace of Mind | • Less convenient than in-person care<br>• Possible improper disclosure of personal information | • Assistance during the pandemic<br>• A good alternative when no other option is available<br>• Independent of transportation<br>• Time and cost saving<br>• Possible improper disclosure of personal information<br>• Rejection by older or technologically challenged individuals | • Assistance during the pandemic<br>• A good alternative when no other option is available<br>• Independent of transportation<br>• Possible improper disclosure of personal information |

### Subtheme: Less Convenient than In-Person Care

Participants identified concerns about the convenience of remote healthcare services compared to in-person visits, especially highlighting the value of physical exams and the personal touch of face-to-face consultations. One participant stated, “In-person visits are more convenient” (IND39). This underscores the interpersonal challenge of remote healthcare services.

### Subtheme: Assistance During the Pandemic

The crucial role of THS services during the pandemic was emphasized, providing necessary support and care in challenging times, significantly when traditional services were disrupted. A participant noted, “It was a great moment. And if you come to apply it in the case of Culebra, it would be phenomenal, too. I would want to have a doctor all the time, but if I have no other option, well, it’s great that this option is there” (IND2). This highlights the community and societal benefits of THS during crises.

### Subtheme: A Good Alternative When No Other Option is Available

Participants viewed THS services as beneficial alternatives when in-person care is inaccessible, mainly due to transportation issues. One participant shared, “I don’t know about anyone else, but I do. I prefer a doctor on camera than none at all” (IND39). This demonstrates the individual, community, and societal advantages of having THS as an option.

### Subtheme: Lack of Knowledge

More knowledge about THS services and procedures still needs to be provided, which may prevent individuals from fully utilizing them. A participant admitted, “I haven’t done it yet” (IND49). This reflects the individual-level challenge of awareness and education regarding THS.

### Subtheme: Independent of Transportation

The advantage of THS services not requiring transportation was particularly noted as useful for communities with limited access to transportation. One participant explained, “In bad weather that there is no transportation which cannot come, then that would be a good alternative” (IND39). This benefit spans community and societal levels.

### Subtheme: Time and Cost Savings

Participants identified significant time and cost savings with the use of remote or THS services, eliminating the need for travel and reducing the opportunity cost of seeking medical care. One participant stated, “These trips would actually waste money and time” (FG103). Another noted, “It saves a lot of time because the kids don’t have to miss school. It’s a whole lost day” (IND1). These points highlight the individual and community benefits of THS.

### Subtheme: Privacy Issues-Improper Disclosure of Personal Information

Concerns were raised about the potential for improper disclosure of personal information when using remote services, which may deter some from utilizing these services. A participant expressed, “There are also many people who do not like to give “[their]” data over the phone. There have been many hackers” (IND49). This concern affects individual, interpersonal, community, and societal levels.

### Subtheme: Rejection by Older or Technologically Challenged Individuals

Participants identified that older individuals or those who are less technologically savvy may resist using remote healthcare services due to unfamiliarity with technology or a preference for traditional in-person care. One participant’s opinion was, “Because sometimes there are many elderly people, they don’t know technology” (IND49). This reflects the individual and community challenges in THS adoption.

### Subtheme: Trust Depends on the Type of Medical Examination

Trust in the THS varies depending on the type of medical examination required. For visual assessments or follow-ups, remote consultations may suffice, but for more tactile, in-depth examinations, in-person visits are preferred to ensure safety and accuracy. One participant remarked, “The decision depends on whether it is a follow-up consultation or if it is the first time with the specialist. If it is an evaluation or, if they need to do a laboratory check, and if they are going to do a study later. Therefore, people should go in person” (FG103). This indicates the individual-level trust considerations.

### Subtheme: Peace of Mind

Having healthcare consultations in the comfort of their own home provides patients with peace of mind. It allows them to communicate more effectively, ensuring they remember to discuss all their concerns with the doctor. A participant shared, “Yes, I sit in my room and explain everything to him. Sometimes, you go to see the doctor, and you forget the things that you had to tell him” (IND59). This benefit is significant at the individual level.

## DISCUSSION

Our findings suggest that the integration of THS with the support of CHWs was perceived positively by the community as an effective approach to improve access to healthcare for those with chronic diseases living in rural settings. Participants indicated that CHWs can positively impact some identified SDoH, such as transportation challenges, particularly on the island, where a golf cart or a clinic official vehicle was proposed to offer patients transportation to and from the clinic. However, it is essential to note that some of these structural determinants include economic instability and geographical isolation, which are beyond the scope of CHWs alone. This study found that the community perceived THS) along with the support of CHWs, as a positive sign to improve access to healthcare. Moreover, participants’ perspectives confirmed that an intervention of CHWs-assisted THS can reduce the lack of specialized healthcare access for patients living in rural settings.

The integration of CHWs and THS can help mitigate barriers related to transportation, provide timely medical consultations, and offer continuous support for managing chronic conditions. The combination of technology and personalized care from CHWs can foster trust and enhance patient engagement, leading to better health outcomes.(26)

The SEM served as a theoretical framework that helped us better understand the impact of interventions using technology, such as THS, whether used alone or assisted by CHWs. SEM allowed for a comprehensive analysis of how individual, interpersonal, community, and societal factors interact to influence health outcomes. We were able to identify people’s perspectives towards the potential integration of CHWs and THS to solve the challenges of healthcare access. Results suggest that CHWs can provide comfort and trust to patients, assist with patient education, schedule medical appointments, manage medications, and offer companionship during medical visits. On the other hand, THS could solve health access challenges, including the time-saving and cost-effectiveness of medical appointments, transportation independence, and a more accessible tool for better chronic disease management. The results confirm that this framework helps to better understand the perspectives and attitudes of people toward integrating CHWs and the THS model for chronic disease management in rural areas.

Findings from the present study confirm participants’ challenges and perceptions toward CHW and THS. We found that these two evidence-based models can be and should be implemented in rural and underserved areas as part of an equitable model for healthcare. However, while CHWs and THS show promise in addressing certain SDoH and improving healthcare access in rural areas, addressing the broader structural determinants requires systemic changes and policy interventions. Future efforts should enhance THS infrastructure, increase community health literacy and willingness, and support CHWs (training) to ensure sustainable and equitable healthcare solutions for rural and isolated populations. By leveraging the strengths of both CHWs and THS, we can move towards a more integrated and effective approach to reducing health disparities and improving the overall health of communities like Culebra.

From a health equity perspective, the findings also inform strategies to address similar issues in other countries, particularly in rural island communities. These communities often face unique challenges such as geographic isolation, limited healthcare resources, and higher vulnerability to health disparities.

Previous studies have shown that CHWs can help navigate health services for Latinx communities who face challenges such as limited resources during COVID-19.(27) Studies have also demonstrated THS as a powerful tool for routine provision of care for Latinos.(28) However, non-previous studies have integrated both CHWs and THS to use them together. By adopting the evidence-based models of CHWs and THS, rural island communities can improve healthcare access, enhance health education, and build sustainable healthcare systems. This can lead to better health outcomes and reduced health disparities, ultimately benefiting communities like Culebra and similar rural island populations globally.

### Study Limitations

The qualitative findings are based on subjective self-reported data, which participants’ personal biases may influence. This study was limited by factors that could influence participant’s perceptions and experiences, including prior healthcare experiences, individual health literacy, and personal attitudes toward technology and healthcare providers.

## Data Availability

All data produced in the present study are available upon reasonable request to the authors

## Acknowledgments

The study team would like to thank the HealthproMed Inc. Culebra clinic staff members for their support during the implementation of the study. In addition, the study team acknowledges Mujeres de Islas Inc. (Culebra community-based organization) for allowing the study team to recruit and promote our study and the Puerto Rico Public Health Trust for CHWs capacity training.

## CRediT Taxonomy

### Contributor Roles

Conceptualization: José G. Pérez-Ramos, Scott McIntosh, Joselyn Wei-Chen

Data curation: José G. Pérez-Ramos, Joselyn Wei-Chen, Jessica Alicea-Vellon

Formal analysis: José G. Pérez-Ramos, Joselyn Wei-Chen

Funding acquisition: José G. Pérez-Ramos

Investigation: José G. Pérez-Ramos, Jessica Alicea-Vellon Methodology: José G. Pérez-Ramos, Scott McIntosh, Joselyn Wei-Chen

Project administration: José G. Pérez-Ramos, Jessica Alicea-Vellon, Joselyn Wei-Chen Resources: José G. Pérez-Ramos, Jessica Alicea-Vellon

Software: José G. Pérez-Ramos, Joselyn Wei-Chen, Jessica Alicea-Vellon Supervision: José G. Pérez-Ramos, Scott McIntosh

Validation: José G. Pérez-Ramos, Scott McIntosh Visualization: José G. Pérez-Ramos, Joselyn Wei-Chen

Writing original draft: José G. Pérez-Ramos, Joselyn Wei-Chen, Scott McIntosh, Carlos Rodriguez-Diaz

Writing review & editing: José G. Pérez-Ramos, Joselyn Wei-Chen, Scott McIntosh, Jessica Alicea- Vellon, Carlos Rodriguez-Diaz

